# Developing a Fully Automated Imaging Biomarker for HCC Risk Assessment via MRI-Based Tumor Segmentation and EPM

**DOI:** 10.1101/2025.11.14.25340275

**Authors:** Awj Twam, Shane A. Smith, Mohamed Eltaher, Zaina Boriek, Dontrey Bourgeois, David Martinus, Tiffany L. Calderone, Laura Beretta, Jessica I. Sanchez, David W. Victor, Nakul Gupta, Manal Hasan, Jalal K. Prasun, Eugene J. Koay, David T. Fuentes

**Author notes:** Corresponding co-first authors.

## Abstract

This study investigates the feasibility of using automated tumor segmentation as the region of interest for early detection of hepatocellular carcinoma (HCC) using magnetic resonance imaging (MRI). Enhancement Pattern Mapping (EPM) is used as a voxel-wise imaging biomarker within the region of interest once segmentations are generated. We implement PocketNet, a lightweight convolutional neural network, to segment liver tumors and evaluate performance. To contextualize model accuracy, we estimate the theoretical upper bound of segmentation performance through inter-observer comparisons of manual annotations. Imaging-derived features from automated segmentations are analyzed using XGBoost to assess their predictive value. Results show that automated segmentation approaches upper-bound performance for larger tumors but underperforms on smaller lesions. However, EPM features derived from automated masks demonstrate comparable predictive power to those from manual segmentations, indicating that the EPM features are stable with respect to segmentation inaccuracies. Results demonstrate that a fully automated imaging biomarker may be developed as a clinical utility for HCC risk assessment.

## INTRODUCTION

Hepatocellular carcinoma (HCC) is the most common primary liver malignancy and a leading cause of cancer-related mortality worldwide.^1^ Early detection through consistent surveillance is essential for improving patient outcomes, particularly among high-risk populations such as those with cirrhosis.^2,3^ Imaging techniques, especially magnetic resonance imaging (MRI), play a pivotal role in HCC detection and characterization, offering superior soft-tissue contrast and functional imaging capabilities.^4^

Manual segmentation is the current gold standard for tumor delineation but is time-consuming and prone to inter-observer variability.^5^ Auto-segmentation models have been developed to streamline lesion segmentation, but their accuracy and reliability compared to manual delineation remain areas of ongoing research. Segmentation performance is known to vary across anatomical sites and tumor sizes. In liver segmentation studies, larger tumors (>3 cm) often yield high Dice scores in the range of 0.85-0.90, whereas small tumors (<1.5 cm) typically achieve Dice scores of only 0.6-0.7, with poor boundary conformity. ^6^ In a recent UNet++□ based segmentation study on liver MRI, the reported DSC for tumors was 0.612 in the validation set and 0.687 in internal testing.^7^ These trends are mirrored in other organ systems: lung nodules^8^, pancreatic tumors^9^, and brain metastases^10^ all exhibit diminished segmentation performance for small lesions. The inherent complexity of liver lesions, coupled with variations in contrast to noise ratio, image quality and tumor heterogeneity, presents significant challenges for automated approaches.^4^ To address the issue of small tumor detection, researchers have introduced multi-scale networks^11^, attention mechanisms^12^, and ensemble strategies to improve sensitivity^13^. Some studies also use radiomics-informed segmentation, where semantic and texture-based priors guide refinement.^14^ One emerging approach for improving tumor detection is Enhancement Pattern Mapping (EPM), a voxel-based image analysis technique that calculates the root mean square difference (RMSD) across imaging to identify tissue at risk of malignant transformation.^15^

Ultimately, tumor segmentation is an intermediate step in the image driven HCC risk assessment workflow. Our approach in this manuscript is to evaluate the sensitivity of HCC risk assessment on tumor segmentation accuracy. This study investigates the feasibility of using automated segmentation masks to drive EPM analysis. Results are compared against manual segmentations as a control. We further explore the upper bound of segmentation performance by benchmarking against inter-observer variability. The concept of an upper bound on segmentation accuracy has been investigated from both theoretical and empirical perspectives. Prior work by Liu and Jha introduced a Cramer-Rao Bound (CRB)-based framework to quantify the theoretical maximum accuracy achievable when ranking quantitate imaging (QI) methods in the absence of a ground truth. Their method estimated the best-case ranking performance of segmentation algorithms under known noise and slope assumptions using statistical bounds.^17^ Complementing this, Udupa et al. Proposed SparseGT, a practical framework for simulating full ground truth segmentation from sparsely annotated slices. By leveraging inter-segmenter variability as a reference, SparseGT established a realistic, empirical ceiling on segmentation accuracy, demonstrating that pseudo-ground truth (p-GT) can achieve human-level performance with up to 96% reduction in manual effort.^18^

Using PocketNet^16^, a lightweight convolutional neural network, we assess the performance of automated tumor delineation and its downstream clinical relevance via EPM-based prediction. Our goal is to establish feasibility of automated imaging biomarkers for HCC detection.

## METHODS

### Dataset and Image Preprocessing

Manual segmentations for tumors were performed by trained experts, establishing the ground truth. The PocketNet architecture was trained on a cohort of 389 images, comprising 261 patients from MD Anderson Cancer Center (MDACC), 46 from Baylor College of Medicine (BCM), and 45 from Houston Methodist Hospital Research Institute. All images were converted to NIfTI format and preprocessed using the Medical Imaging Segmentation Toolkit (MIST, version 0.4.1b). Preprocessing steps included resampling to uniform voxel spacing (0.742 x 0.742 x 2.0 mm), reorientation to RAI, and Z-score intensity normalization based on non-zero voxels. Training data used a patch size of 256 x 128 x 64, derived from the median resampled image dimensions.

### Network architecture and Training Protocol

Tumor segmentation was performed using PocketNet, a lightweight convolutional neural network optimized for 3D medical image segmentation.

The PocketNet architecture was implemented following the methodology outlined in the cited work.^16^ Training was conducted using an Nvidia Quadro RTX 8000 GPU. The model was trained for 1,000 epochs using the ADAM optimizer. Oversampling rate was set to 1.0. Loss was computed using a weighted combination of Dice and categorical cross-entropy. Automated mixed precision (AMP) was used to accelerate training.

### Performance Metrics

Segmentation accuracy was assessed using the Dice Similarity Coefficient (DSC) and the 95th percentile Hausdorff distance (Haus95) to evaluate spatial agreement and boundary accuracy.

### Upper Bound Analysis

To contextualize automated segmentation performance, we estimated an upper bound using inter-observer variability from a subset of cases (Methodist, N=45) with multiple manual annotations. Specifically, Dice Similarity Coefficient (DSC) values were computed pairwise between each pair of expert raters. The average of these pairwise DSCs reflects the typical level of agreement achievable between human annotators and thus serves as a practical ceiling or upper bound for segmentation performance.

This empirical ceiling serves as a benchmark for evaluating automated models, indicating the maximum feasible performance under real-world constraints. Together, these metrics provide a multifaceted view of performance limitations, guiding interpretation of results, and the feasibility of future improvements.

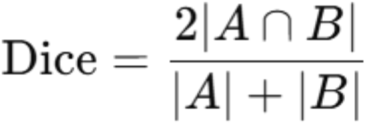

**Equation 1)** Pairwise DSC calculation, where A represents the predicted tumor mask and B represents the ground truth mask.

### HCC risk analysis

To evaluate clinical relevance beyond segmentation, we constructed a risk classification dataset derived from a cohort of patients from BCM with cirrhosis undergoing serial multiphase contrast-enhanced MRI as part of a prospective HCC surveillance protocol.^19^ Each patient had imaging data at two key clinical stages: a diagnostic (DX) timepoint (when HCC was radiologically or pathologically confirmed) and a prediagnostic (PX) surveillance scan preceding diagnosis by ∼6 months. Controls (DX n=100, PX n=93) included patients with cirrhosis, with or without indeterminate liver lesions (LI-RADS 3 or 4), who did not progress to HCC. Cases (DX n=48, PX n=47) were defined as patients who had a LI-RADS 3 or 4 lesion identified on a pre-diagnostic scan that subsequently progressed to HCC on a diagnostic scan. After excluding patients for poor image quality (motion artifact) or unavailable imaging, the final dataset comprised 148 total eligible patients.

At both timepoints, automated segmentations were obtained for cases and controls with EPM and shape features being obtained for each patient at both timepoints; the features measured are described in Table First order and shape metrics ROIs were obtained segmented liver regions from automated masks. Distance metrics were calculated between the voxel distributions of automated mask ROIs and non-segmented background liver. Values for shape features are calculated in a manner that weighs larger ROIs more heavily if multiple ROIs are present on an automated segmentation. A full breakdown of shape and distance calculations is provided in the supplementary data.

### Model Training and Feature Selection

Using our EPM and shape-based feature dataset, we performed binary classification via XGBoost to determine if automated segmentations could distinguish between cases from controls at each timepoint. Model evaluation was performed using 5-fold stratified cross-validation with performance being measured using accuracy, precision, recall, and F1-score. A full breakdown of model training, feature selection, and hyperparameter selection is provided in the supplementary data. Additionally, we also tested classification with other common models such as Logistic Regression, Support Vector Machine, Random Forest, Decision Tree, and K-Nearest Classification. Results of these models are also provided in supplementary data.

### Ethics Approval

This study was approved by the Institutional Review Boards (IRB) of The University of Texas MD Anderson Cancer Center (Protocol No. [PA15-0091]), Houston Methodist Research Institute (Protocol No. [PA13-0317]), and Baylor College of Medicine (Protocol No. [PA14-0646]). The requirement for informed consent was waived by these IRBs due to the retrospective nature of the study and the use of de-identified data. All experiments and analyses were conducted in accordance with institutional guidelines and regulations.

## RESULTS

### Automated Tumor Segmentation Performance Analysis

We first evaluated the performance of the PocketNet model on the full liver tumor training dataset (N=397), which included cases from multiple institutions. The performance of the PocketNet architecture revealed a mean DSC of 0.52 (standard deviation:0.28, median: 0.58, 75^th^ percentile: 0.74), indicating a modest level of accuracy in automated liver tumor segmentation.

To illustrate segmentation variability, Figure 3 presents three cases demonstrating the performance of the PocketNet architecture relative to manual tumor segmentation. DSCs, indicating spatial overlap and segmentation accuracy, are shown for each case ranging from 0 (no overlap) to 1 (perfect overlap). The top row (DSC: 0.83) represents a high concordance between manual and automated segmentation, indicating effective automated delineation. The middle row (DSC: 0.54) illustrates moderate segmentation accuracy. The automated segmentation captures the tumor location adequately but differs notably from the manual standard, indicating challenges in precise replication of manual contours. The bottom row (DSC: 0.37) demonstrates a lower agreement, highlighting significant discrepancies between automated and manual segmentations.

**Figure 1).**
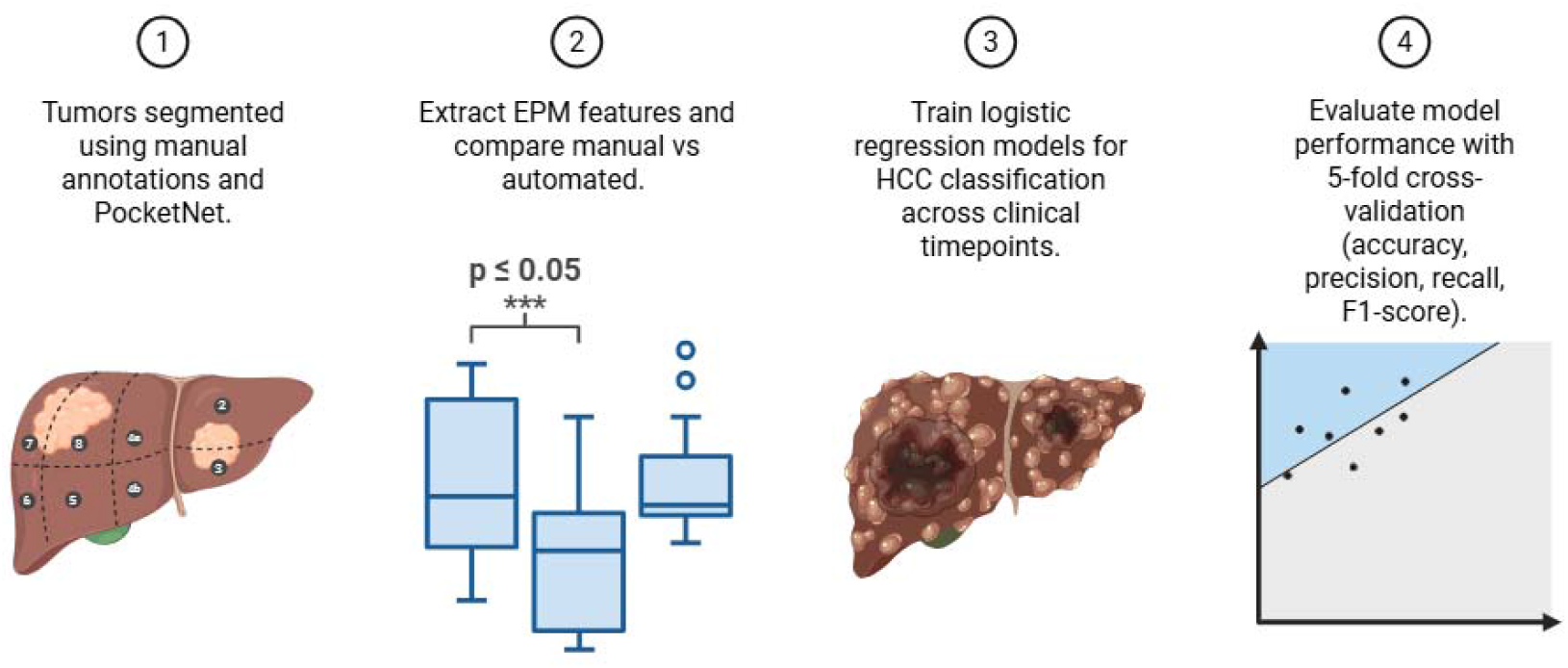
Workflow for segmentation and risk prediction analysis. Automated (PocketNet) and manual tumor sementations were compared as inputs for EPM, shape, and distance feature extraction. A subset of patients with complete imaging across clinical timepoints (PX, DX) was used for downstream risk classification. XGBoost models were trained to distinguish cases from controls with performance being evaluated with stratified 5-fold cross-validation using per-class accuracy, precision, recall, and F1 scores.

**Figure 2).**
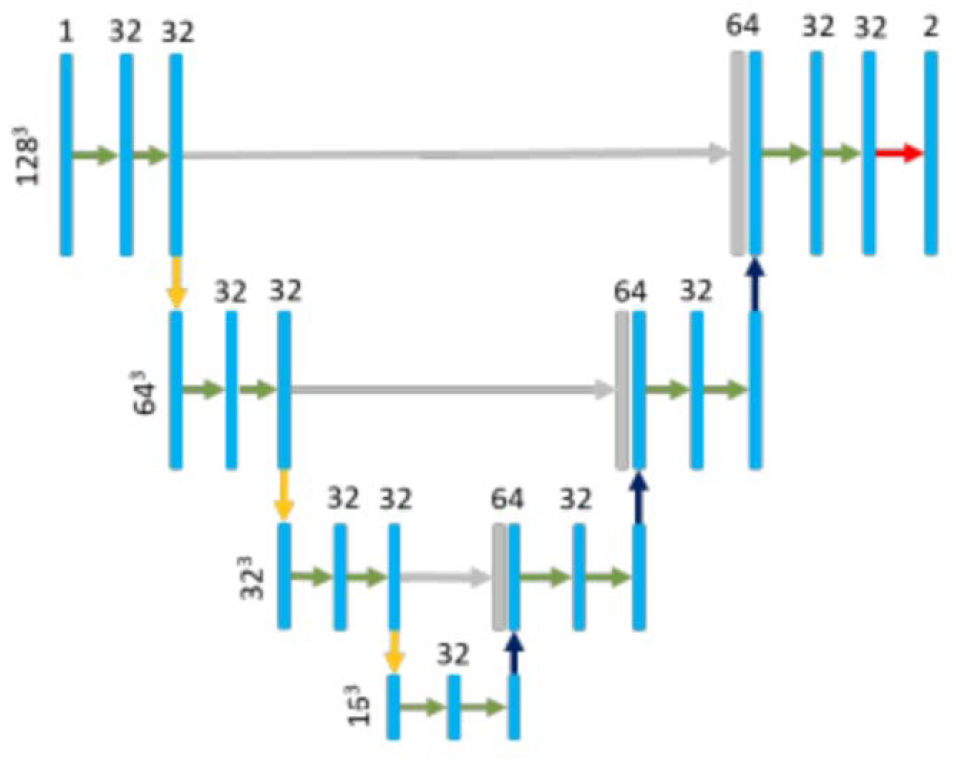
*PocketNet* Architecture, a compact convolutional neural network designed for medical image segmentation.

**Figure 3).**
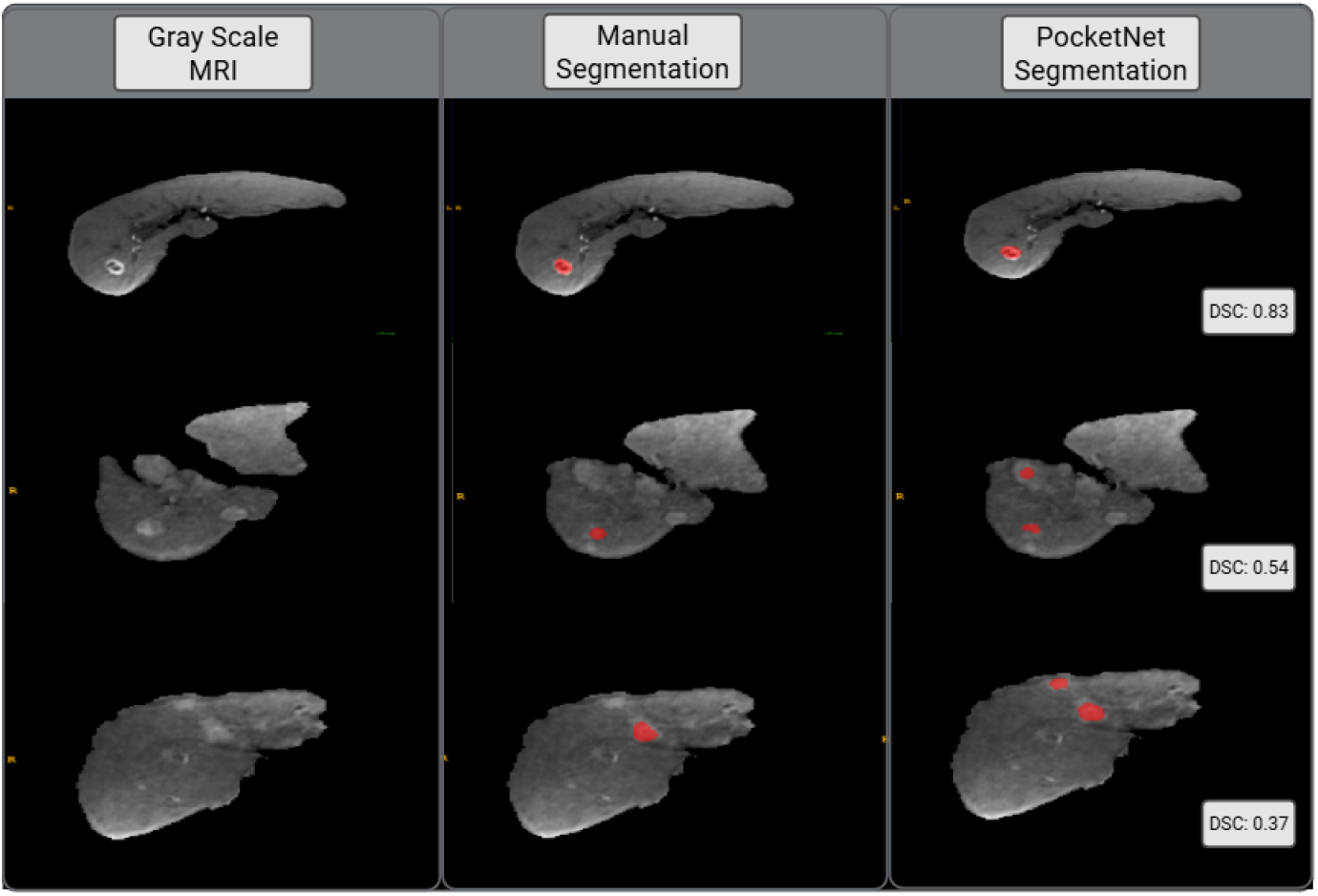
Comparison of Manual and Automated (PocketNet) Tumor Segmentation at Varying DSC Scores. Each row depicts a separate patient case, displaying axial MRI slices. The left column represents a gray-scale T1 axial MR image of the liver. The middle column (“Manual Segmentation”) shows tumor contours manually delineated by experts, while the right column (“PocketNet Segmentation”) shows the corresponding contours predicted by the automated algorithm.

### Upper Bound Estimation for Segmentation Accuracy

To investigate the automated segmentation performance, we evaluated a subset of the dataset (Methodist cohort, N=45) where tumor segmentations from three independent manual raters were available. This subset is the only portion of the dataset with multiple annotations, allowing us to assess inter-observer variability as an empirical upper bound for achievable segmentation accuracy.

Pairwise comparisons between the three raters yielded a mean DSC of 0.62, suggesting a moderate level of agreement among annotators. We denote these comparisons as DSC1, DSC2, and DSC3 representing:

- DSC1: Rater A vs Rater B
- DSC2: Rater B vs Rater C
- DSC3: Rater A vs Rater C

These comparisons represent human-level segmentation variability and serve as a reference ceiling against which automated results can be evaluated. Mean DSC values between the automated model and each manual rater were also computed and overlaid on the plots as a red line, allowing for a direct visual comparison between human and algorithmic performance. The close alignment between the red line and the medians of the manual distributions illustrates that the automated model performs within the general range of human annotators, though it remains slightly below expert-level agreement. The presence of outliers at 0 indicates some cases of significant disagreement, highlighting the challenges in consistently defining tumor boundaries. These variations underscore the complexity of manual segmentation and the critical need for more robust automated approaches.

To further evaluate the agreement between automated and manual tumor segmentation, we performed a Bland-Altman analysis using the same Methodist cohort of 45 cases. This analysis provides a more granular comparison between automated and manual segmentations in a context where inter-observer variability can also be assessed.

The Bland-Altman plot in Figure 5 illustrates the agreement between the automated and manual segmentation of liver tumors in the Methodist subset. This graphical analysis assesses the degree of concordance between two segmentation methods by plotting the differences between paired Dice Scores (automated minus manual) against their mean dice scores. The central dashed line on the plot represents the average difference between automated and manual segmentation, calculated as –0.104. This negative mean difference indicates that, on average, the automated segmentation yields slightly lower Dice scores compared to manual segmentation.

**Figure 4).**
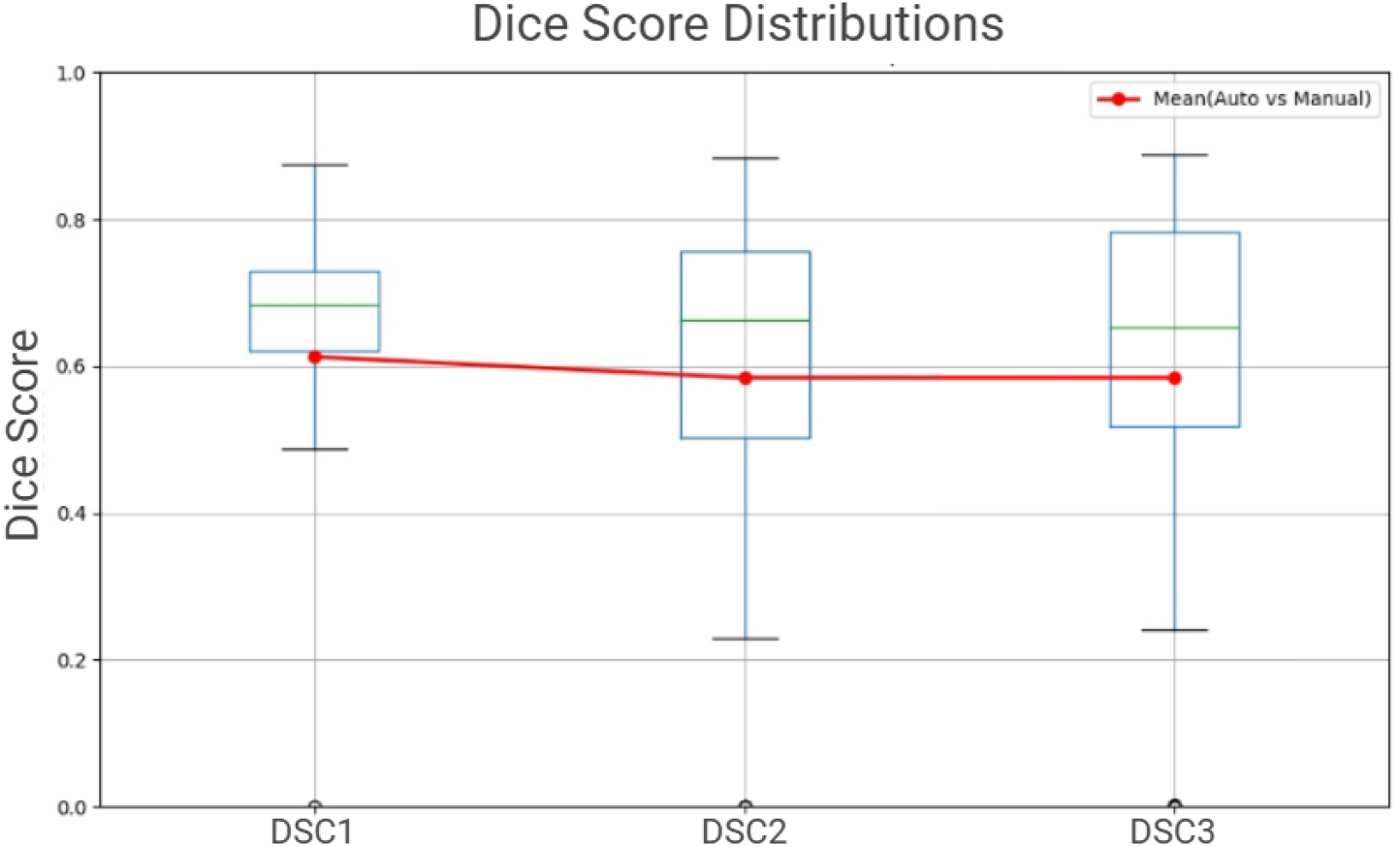
Dice Score distributions for manual segmentation comparisons (DSC1, DSC2, DSC3), representing inter-observer variability on the Methodist subset. Red dots and the connecting line indicate the mean Dice scores between the automated segmentation and the consensus of each rater pair.

**Figure 5).**
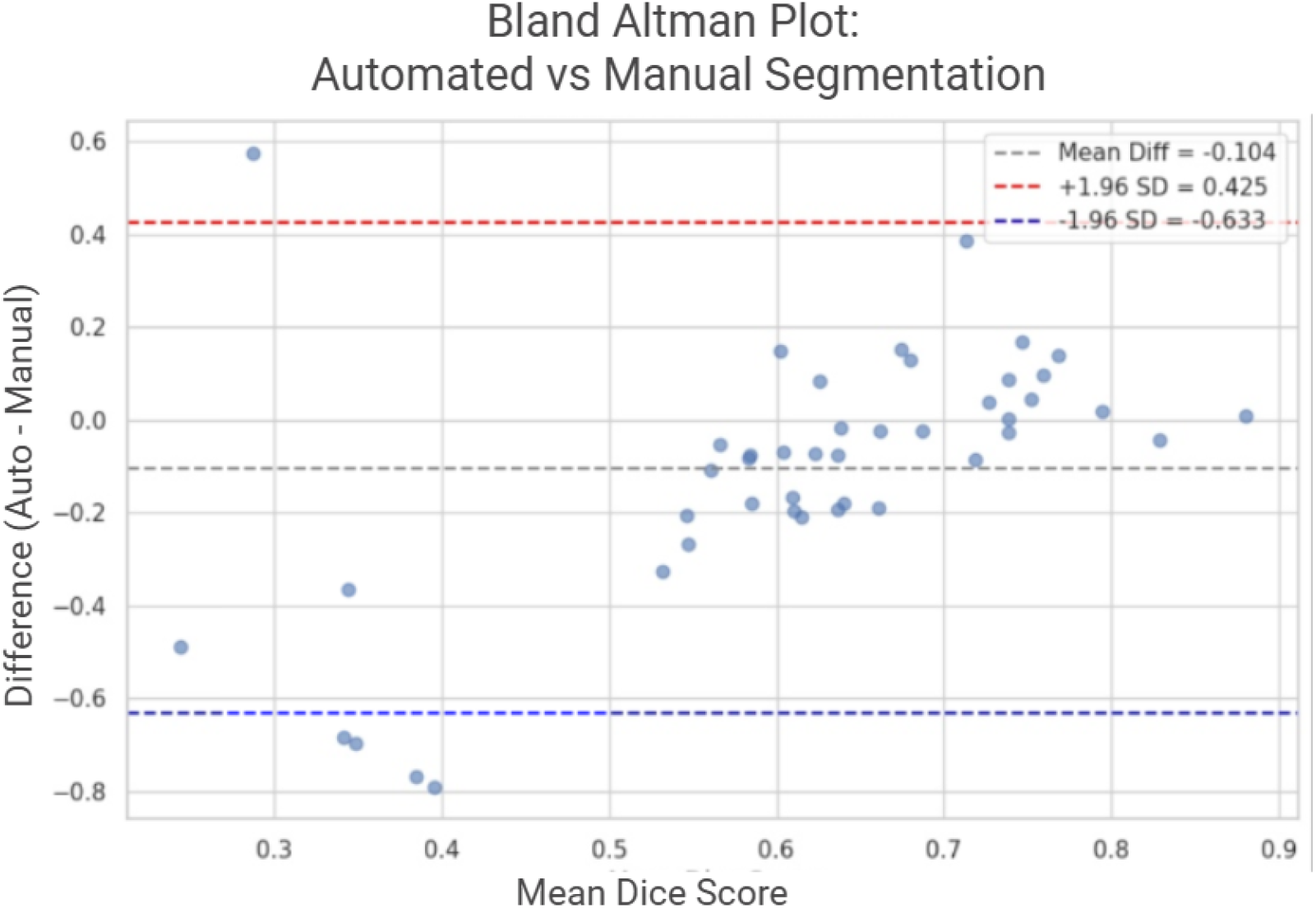
Bland-Altman Analysis of Agreement: Automated vs Manual Tumor Segmentations (Methodist subset).

The upper limit of agreement (+1.96 standard deviations) is marked by the red dashed line at approximately 0.425, while the lower limit (-1.96 SD) was -0.633, with a standard deviation of 0.2698. These values define the range within which most differences between automated and manual DSC values lie. The limits of agreement reflect the variability in overlap between the two methods, with clustering near the mean difference line indicating moderate agreement overall and points near the extremes representing cases of divergence.

### Predictive Utility via Binary Classification

Building on the segmentation accuracy results, we next examined whether automated tumor masks capture the same quantitative lesion characteristics as expert manual annotations and thus could determine true HCC cases from controls. Separate XGBoost models were trained for each timepoint: diagnostic and prediagnostic. For each model, EPM summary first order statistics, distance metrics, and shape metrics from automated segmentations served as predictors.

Summaries of model performances are given in Tables 2 and 3 as well as Figures 6, 7, and 8. Our findings suggest that imaging-based features extracted from automated tumor masks are reliable and able to differentiate true HCC lesions from non-HCC control segmentations at crucial diagnostic timepoints. Additionally, these imaging-based features from automated tumor masks may provide meaningful information even before radiologic evidence of HCC becomes apparent, but the extent of this prognostic value may be limited. These results collectively demonstrate that, despite modest segmentation accuracy, the automated outputs still retain clinically meaningful signal.

**Table 2)).** Summary of evaluated metrics: first order, distances, and shape metrics at diagnostic and prediagnostic timepoints.

| METRIC<br>EVALUATED | DESCRIPTION<br>OF METRIC | CASE-CTRL<br>P-VALUE<br>(DX \| PX) |
| --- | --- | --- |
| **FIRST ORDER** |  |  |
| Mean | Average intensity value within lesion ROI – reflects overall enhancement magnitude | 0.44 \| 0.05 |
| Mode | Most frequently occurring intensity value in lesion ROI | <0.01 \| <0.01 |
| Median | Middle intensity value within lesion ROI. Less sensitive to outliers than mean | 0.63 \| <0.01 |
| Standard Dev. | Square root of variance; measure of intensity dispersion | <0.01 \| 0.13 |
| Variance | Spread of intensity values; indicates heterogeneity of enhancement | <0.01 \| 0.13 |
| Skewness | Asymmetry of intensity values in lesion ROI voxel distribution | 0.15 \| <0.01 |
| Kurtosis | “Tailedness” or extremity of enhancement values; related to prevalence of outliers. | 0.16 \| 0.08 |
| **FIRST ORDER RATIO** |  |  |
| Ratio(Stat) | Ratio of (Lesion Stat)/(Non-Lesion Liver Stat). Measures the | Various |

ratio of ROI intensity to non-ROI liver intensity. Values closest to 1 indicate less difference between ROI and liver.
| DISTANCES |  |  |
| --- | --- | --- |
| Hellinger | Quantifies similarity of two probability distributions – related to Bhattacharyya distance, but unlike this distance, is a true metric | 0.10 \| <0.01 |
| Bhattacharyya | Quantifies similarity of two probability distributions. Not a true metric as the triangle inequality is not obeyed. | 0.08 \| <0.01 |
| Total Variation | Quantifies similarity of two probability distributions. Can represent max bias of an event under two distributions. | 0.33 \| <0.01 |
| Energy | Measure of dissimilarity of two probability distributions; inspired by Newton’s gravitational potential energy. | 0.58 \| <0.01 |
| Earth Mover’s | Represents the minimum “work” or “cost” to transform one probability distribution to another. | 0.33 \| <0.01 |
| SHAPE METRICS |  |  |
| Elongation | Quantifies the linearity or “stretched” nature of an ROI | 0.03 \| 0.75 |
| Flatness | Defines how flat, non-spherical an object is. | 0.01 \| 0.87 |
| Least Axis Length | Smallest axis length of the ROI-enclosing ellipsoid | <0.01 \| 0.11 |
| Minor Axis Length | Second-largest length of the ROI-enclosing ellipsoid | <0.01 \| 0.12 |
| Major Axis Length | Largest axis length of the ROI-enclosing ellipsoid | <0.01 \| 0.12 |
| Sphericity | Roundness of the shape of the ROI region relative to a sphere | 0.85 \| 0.91 |
| Surface Area | Calculated surface area of the ROI | <0.01 \| 0.10 |
| Surface-Volume Ratio | Ratio of ROI surface area to volume – lower values typically indicate more compact objects. | <0.01 \| 0.20 |

**Table 2).**
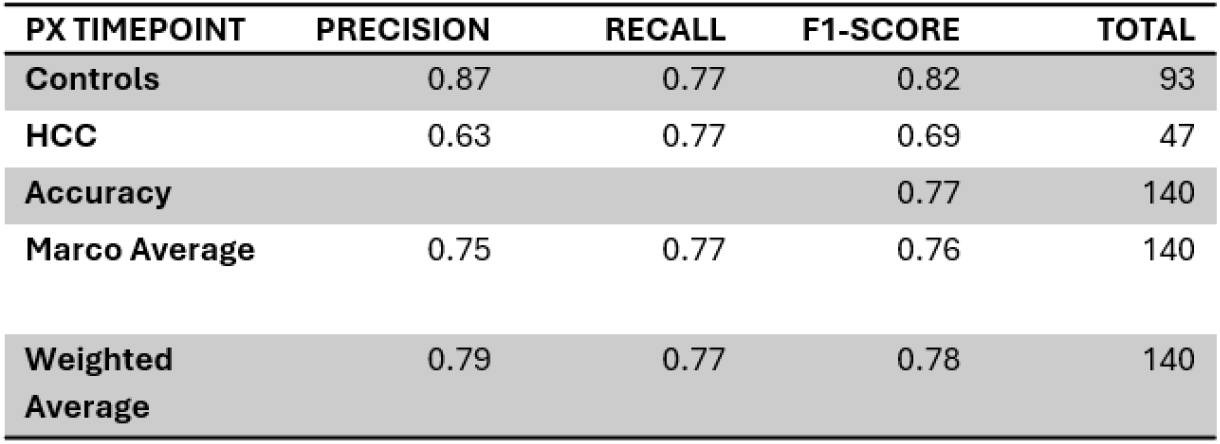
Classification report of binary classification via XGBoost at the prediagnostic timepoint.

| PX TIMEPOINT | PRECISION | RECALL | F1-SCORE | TOTAL |
| --- | --- | --- | --- | --- |
| Controls | 0.87 | 0.77 | 0.82 | 93 |
| HCC | 0.63 | 0.77 | 0.69 | 47 |
| Accuracy |  |  | 0.77 | 140 |
| Marco Average | 0.75 | 0.77 | 0.76 | 140 |
| Weighted Average | 0.79 | 0.77 | 0.78 | 140 |

**Table 3).** Classification report of binary classification via XGBoost at the diagnostic timepoint.

| DX TIMEPOINT | PRECISION | RECALL | F1-SCORE | TOTAL |
| --- | --- | --- | --- | --- |
| Controls | 0.93 | 0.90 | 0.91 | 100 |
| HCC | 0.80 | 0.85 | 0.83 | 48 |
| Accuracy |  |  | 0.89 | 148 |
| Marco Average | 0.87 | 0.88 | 0.87 | 148 |
| Weighted Average | 0.89 | 0.89 | 0.89 | 148 |

**Figure 6).**
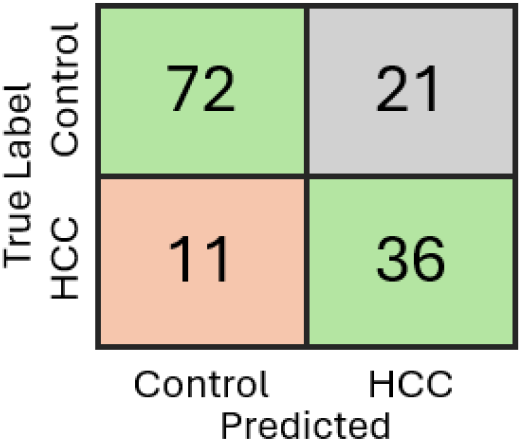
Confusion matrix of binary classification via XGBoost at the prediagnostic timepoint.

**Figure 7).**
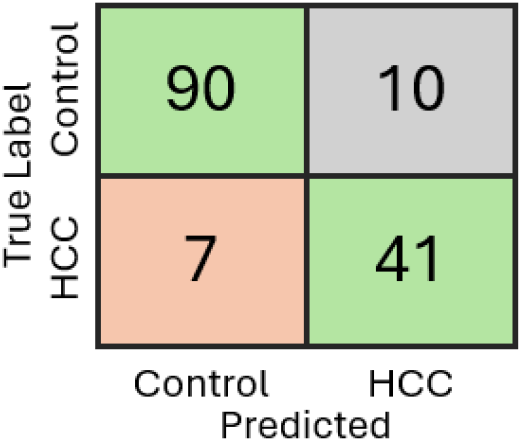
Confusion matrix of binary classification via XGBoost at the diagnostic timepoint.

**Figure 8).**
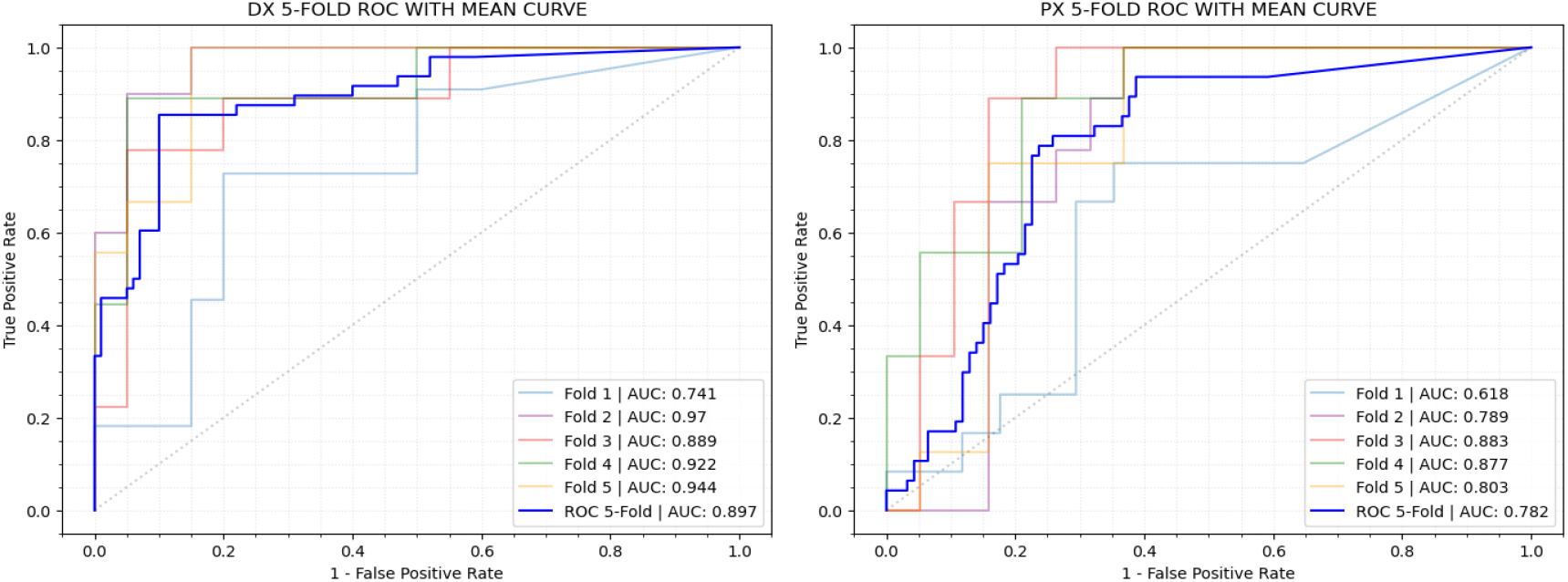
ROC analysis of XGBoost models when stratified 5-fold cross-validation is applied. ROC curves are shown with AUC for each individual fold, as well as an aggregate ROC curve representing the overall performance at **(A)** the diagnostic timepoint and **(B)** the prediagnostic timepoint.

## DISCUSSION

This study evaluated the feasibility of using automated liver tumor segmentation generated by the PocketNet architecture as the basis for extracting a comprehensive set of imaging biomarkers-including EPM, shape, and distance-based features to predict HCC risk in patients with cirrhosis. By combining conventional spatial accuracy metrics with inter-observer upper bound analysis and downstream predictive modeling, we provide a multidimensional view of algorithm performance and clinical relevance.

The average DSC of 0.52 achieved by our PocketNet architecture suggests that current automated methods still face limitations in accurately delineating tumor boundaries. While this may appear modest, it must be contextualized within the inherent variability of manual segmentation itself. Our upper bound analysis, based on inter-observer comparisons, demonstrated that even expert annotators rarely agree perfectly, with a mean DSC of 0.62. This finding is crucial for setting realistic expectations for automated approaches and highlights the subjective nature of tumor boundary delineation. The 0.52 accuracy rate does not necessarily represent a complete failure of the approach, but rather a starting point for further methodological development. It reveals the intricate challenges in translating deep learning techniques to the nuanced domain of medical image analysis. Factors such as image quality, tumor heterogeneity, and the inherent complexity of liver lesions likely contribute to the current limitations.

The Bland-Altman analysis further illustrated agreement patterns between manual and automated segmentations within the Methodist subset. The analysis revealed a small systematic bias, with automated segmentations tending to achieve slightly lower Dice scores than manual references, and moderate variability across cases. The positive trend observed in the plot suggests that agreement between manual and automated segmentations improves for cases with higher mean Dice scores, implying that the model performs best for larger or more conspicuous legions.

The predictive modeling results further support the utility of these features. XGBoost classification models revealed that our automated segmentations can achieve modest to high accuracies in classifying cases and controls, indicating that the predictive capabilities of imaging-derived features are robust at diagnostic and prediagnostic surveillance timepoints. The ability of imaging-derived features from automated masks to predict case status suggests their potential as an early biomarker for patient risk stratification. These findings align with recent advances in radiomic-clinical integration for HCC prognosis. Vithayathil et al. (2025) demonstrated that a deep learning model combining radiomic and clinical features from pretreatment computed tomography (CT) scans of patients receiving atezolizumab plus bevacizumab achieved strong predictive performance (AUC = 0.89) for overall survival and immunotherapy response, surpassing traditional clinical scores.^21^ Their results underscore the translational potential of imaging-derived biomarkers when coupled with machine learning frameworks for individualized prognostication.

Similarly, Li et al. (2025) developed machine-learning based HCC-risk models in patients with HBV-related compensated advanced chronic liver disease, where random-forest and XGBoost algorithms achieved AUCs > .0.94 using clinical and biochemical predictors such as liver stiffness, platelets, bile acid, and white-blood-cell count.^22^ Our feature-driven framework parallels these principles by incorporating geometric and spatial metrics derived from imaging data rather than laboratory features. The results of both studies underscore the translational potential of imaging-derived biomarkers when coupled with machine learning frameworks for individualized prognostication.

Together, these findings offer compelling support for the integration of automated segmentation into the clinical pipelines-not necessarily as a replacement for manual annotation, but as a scalable and reproducible tool for longitudinal surveillance, risk stratification, and early detection. While spatial overlap with expert contours may be moderate, the clinical signal preserved in these segmentations is strong enough to support downstream decision-making.

## CONCLUSION

In this study, we established a fully automated pipeline for EPM-based HCC characterization, incorporating automated tumor segmentation, quantitative feature extraction, and predictive modeling. While automated segmentation achieved a modest DSC of 0.52, the upper bound analysis revealed that even manual segmentations by experts show variability. Crucially, despite imperfect spatial overlap, automated segmentation preserved most of the lesion-level EPM characteristics, enabling XGBoost classification models to achieve up to 89% classification accuracy for distinguishing HCC cases from controls.

Our findings suggest that automated segmentation, even at current performance levels, holds promise for scalable, reproducible risk assessment in HCC surveillance. Future work should focus on improving segmentation accuracy for small and heterogenous lesions, integrating uncertainty estimation to identify low-confidence cases, and validating predictive performance in larger, mult-institutional cohorts with standardized imaging protocols. Such advances could enable deployment of a fully automated EPM-based pipeline for noninvasive, early risk assessment in patients with cirrhosis.

## Data Availability

The patient imaging data used in this study are not publicly available due to institutional privacy policies. However, the analysis and model implementation code used in this study are available upon request.

## Acknowledgements

The authors would like to acknowledge the support of the Tumor Measurement Initiative through the MD Anderson Strategic Research Initiative Development (STRIDE), QIAC partnership in research, CPRIT RP250154, NIH R01CA195524, and R01CA301555.

## Additional Information

### Author Contributions

A.T. and S.A.S. contributed equally to this work and share first authorship. They jointly performed data curation, model development, statistical analysis, and manuscript writing. M.E. performed segmentation of institutional data. D.B. processed and generated the EPM images. T.L.C., L.B., and J.I.S. contributed to data collection and storage. N.G. provided expert radiologic oversight and validation of imaging data. D.W.V., E.J.K., J.K.P., and M.H. contributed clinical expertise, data acquisition, interpretation of imaging and patient data within the study context. D.T.F. supervised the overall study and provided critical feedback. All authors reviewed and approved the submitted manuscript.

