## Supplementary Material for "Developing a Fully Automated Imaging Biomarker for HCC Risk Assessment via MRI-Based Tumor Segmentation and EPM"

### SUPPLEMENTARY DATA

#### MODEL TRAINING, TUNING, AND FUTURE SELECTION

##### Handling Missing Data (Blank Segmentations)

Model training and selection was conducted using a scikit-learn pipeline for logistic regression, support vector machine (SVM), random forest, decision tree, k-nearest, and XGBoost classification models. While more advanced models, such as the XGBoost models described in this study, are capable of handling missing data natively, not all models are capable of this. Simpler models, such as logistic regression, fail in the presence of missing data; this posed a challenge in model training as many control automated segmentations were blank with no detected ROI. As such, all models (even those with missing data support) were trained on the subset of data which did *not* include blank segmentations. This method was selected for training as it avoids potentially altering or masking correlations necessary for accurate classification that would otherwise result from imputation or simply setting missing values to 0.

All models were evaluated on all data, including instances missing data using a stratified 5-fold method that ensured even splits of non-blank cases, non-blank controls, and mixed blanks instances across all folds. In evaluating models, each instance was assigned a binary non-blank coefficient $\beta_{nb}$ that was given a value of 1 if a non-blank automated segmentation was present and 0 if the segmentation was blank. In evaluation, a model's probability output was multiplied by this coefficient to give the final prediction probability; the rationale behind this is that blank segmentations should always be classified as controls since no lesions are present that would indicate HCC while also leaving non-blank probabilities unaltered and able to be used in prediction normally. This modification to the probability prediction is illustrated below with the linear prediction and sigmoid transformation of a logistic regression model. During evaluations, blank statistics were filled with placeholder values of 0 in models not permitting missing values, though this served simply to avoid a coding error – mathematically, the non-blank coefficient allows for 0 probability predictions on blank segmentations while allowing for a fundamentally more stable training on non-blank instances.

$$P(y=1|x) = \beta_{nb}\left(\frac{1}{1+e^{-z}}\right) \text{ (S1)}$$

$$z = \beta_0 + \beta_1 \cdot x_1 + \beta_2 \cdot x_2 + \cdots + \beta_k \cdot x_k \text{ (S2)}$$

**Equation S1)** The probability output of logistic regression featuring the non-blank coefficient $\beta_{nb}$.
**Equation S2)** Unaltered linear predictor of a logistic regression model used in finding probability.

##### Model Feature Selection

Model features were selected by default using Scikit-Learn's Recursive Feature Elimination with stratified 5-fold cross validation at each step (RFECV). This initially fits a model to all available features, evaluates the model, and prunes the least important feature – this process is repeated until the optimized model is found, pruning futures one at a time. Feature importances are obtained on a by-model basis (beta coefficients in logistic regression, contribution of a feature to reducing impurities in decision trees, etc.). Due to limitations in patient counts for the event-positive HCC class, only the best 6 features from RFECV were kept if more than 6 features were returned. By default, RFECV attempts to select features based on a model feature importances in maximizing a model's accuracy, but due to imbalanced case and control counts in this study, this was altered to maximize the model's overall F1 score.

##### Model Hyperparameter Tuning

Model hyperparameters were found using Scikit-Learn's Grid Search with stratified 5-fold cross-validation (GSCV) using models fitted to the found optimal features from RFECV. GSCV brute-force searches for the best parameter combinations for a given model type which optimizes model performance. In this study, that model performance was changed from default accuracy to the F1 score for the same reason RFECV was set to this metric. For each parameter combination, stratified 5-fold cross validation is performed to evaluate model performance given set of parameter combinations. The combination of best parameters is then returned to be used in final model evaluation.

**Model Performance Evaluation**

Performance of models was evaluated using stratified 5-fold cross validation that ensured even splits of non-blank controls, non-blank cases, and mixed blanks across all folds. Models were scored using precision, recall, and F1 score futures for both the case and control classes, with overall model accuracy being taken as well. Confusion matrices and ROC curves are created as well. For ROC curves, a curve is produced for each induvial fold along with a mean curve.  Area under the curve (AUC) is taken for ROC evaluation.

**Model Results**

An overview of model results can be found in Table S1. All models tend to show comparable performance to the best XGBoost models which were described in this study, with the key exceptions being K-Nearest and SVM classifiers which lacked RFECV compatibility.

| MODEL TYPE | HCC PRECISION | CONTROL PRECISION | HCC RECALL | CONTROL RECALL | HCC F1 -SCORE | CONTROL F1-SCORE | MODEL MARCO AVERAGE | MODEL WEIGHTED AVERAGE | MODEL ACCURACY |
|---|---|---|---|---|---|---|---|---|---|
| LOGISTIC REGRESSION, DX | 0.70 | 0.91 | 0.83 | 0.83 | 0.76 | 0.87 | 0.82 | 0.83 | 0.83 |
| LOGISTIC REGRESSION, PX | 0.62 | 0.82 | 0.64 | 0.81 | 0.63 | 0.81 | 0.72 | 0.75 | 0.75 |
| SUPPORT VECTOR MACHINE, DX | 0.60 | 0.80 | 0.58 | 0.81 | 0.59 | 0.81 | 0.70 | 0.74 | 0.74 |
| SUPPORT VECTOR MACHINE, PX | 0.58 | 0.77 | 0.51 | 0.81 | 0.54 | 0.79 | 0.66 | 0.70 | 0.71 |
| DECISION TREE, DX | 0.71 | 0.87 | 0.73 | 0.86 | 0.72 | 0.86 | 0.79 | 0.82 | 0.82 |
| DECISION TREE, PX | 0.59 | 0.88 | 0.81 | 0.72 | 0.68 | 0.79 | 0.74 | 0.76 | 0.75 |
| RANDOM FOREST, DX | 0.80 | 0.88 | 0.73 | 0.91 | 0.76 | 0.89 | 0.83 | 0.85 | 0.85 |
| RANDOM FOREST, PX | 0.60 | 0.84 | 0.70 | 0.76 | 0.65 | 0.80 | 0.72 | 0.75 | 0.74 |
| K-NEAREST CLASSIFIER, DX | 0.54 | 0.78 | 0.54 | 0.78 | 0.54 | 0.78 | 0.66 | 0.70 | 0.70 |
| K-NEAREST CLASSIFIER, PX | 0.55 | 0.76 | 0.51 | 0.78 | 0.53 | 0.77 | 0.65 | 0.69 | 0.69 |
| XGBOOST, DX | 0.80 | 0.93 | 0.85 | 0.90 | 0.83 | 0.91 | 0.87 | 0.89 | 0.89 |
| XGBOOST, PX | 0.63 | 0.87 | 0.77 | 0.77 | 0.69 | 0.82 | 0.76 | 0.78 | 0.77 |

**Table S1)** Precision, recall, F1, and accuracy scores of the models trained and evaluated. Statistics are given in a class-based (HCC-control) format. Highlighted XGBoost models illustrate the models described in this study.

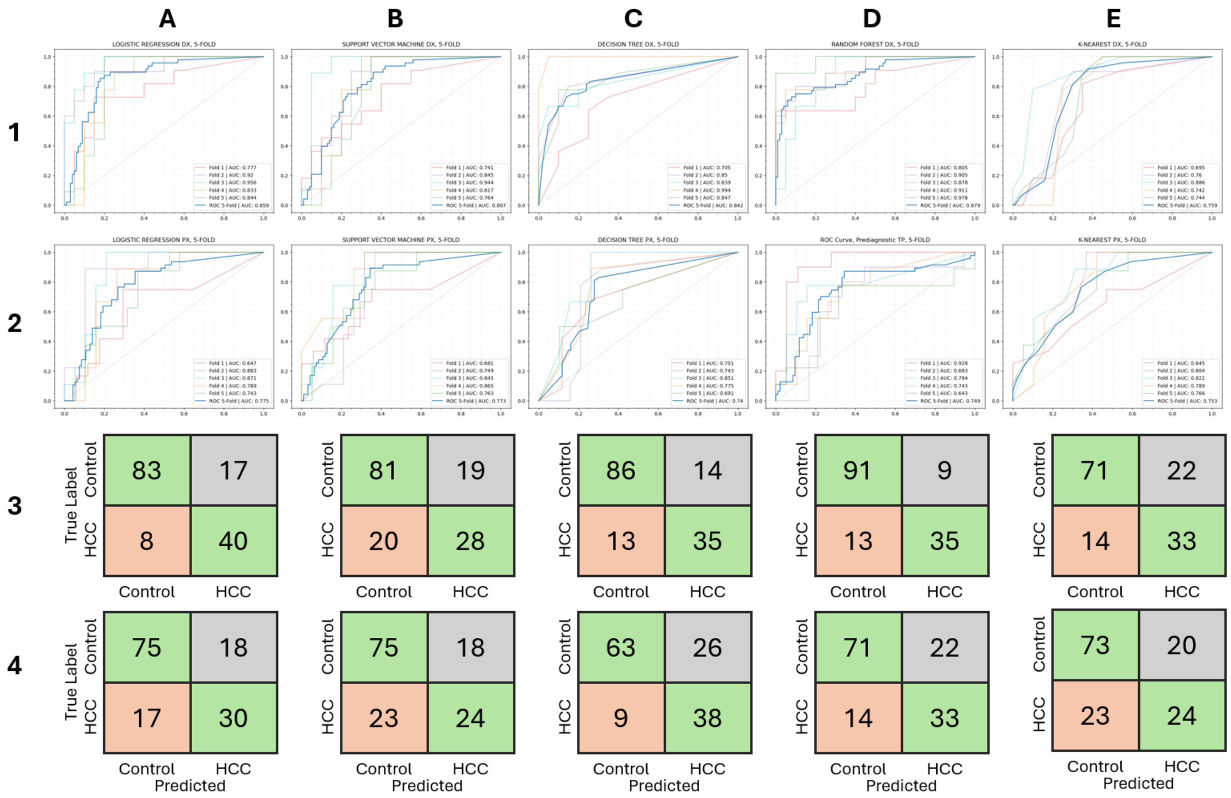


**Figure S1)** ROC curves and confusion matrices for non-XGBoost classification models trained and evaluated. Column **A** contains figures for logistic regression, column **B** contains figures for support vector machine, column **C** contains figures for decision tree, column **D** contains figures for random forest, and column **E** contains figures for K-nearest classifier. For a given model **1** indicates the DX timepoint ROC curve, row **2** indicates the PX timepoint ROC curve, row **3** indicates the DX model confusion matrix, and row **4** indicates the PX model confusion matrix.

#### STATISTICAL DISTANCE CALCULATIONS

##### Implementation of a Gaussian KDE for Liver and ROI distribution Probability Density Functions

Statistical distance metrics were included in this study to evaluate the overall difference between segmented liver regions (supposedly representing lesions) and non-segmented background liver with the goal of evaluating two key hypotheses: 1) segmented HCC lesions will show modest statistical distance from background liver and 2) ROIs segmented on controls will generally show different distance signals than HCC ROIs and thus represent some type of discernable non-HCC noise. For the distances selected in this study, raw observed data must be transformed to its probability density function (PDF) which represent the relative likelihood of a continuous random variable taking on a given value (or in the context of this study, the probability of a voxel intensity occurring given a sample distribution of all voxel intensities). Alternatively, summation-based equations also exist for these distances which instead use the two discrete probability distributions from observed data; this method is typically preferred when observed data is used. In the context of this study however, a major challenge in taking these distances using discrete probability distributions is that summation-based equations require discrete probability distributions to contain the same amount of samples; given that non-segmented background liver distributions are significantly larger than distributions of segmented ROIs (i.e. a >100,000 voxel liver distribution versus a 400-1000 voxel lesion distribution), this all but guarantees the two discrete probability distributions will be vastly different in size.

While resampling of either discrete probability distribution to match the size of the other is technically possibly, the sheer risks of data loss or artifact introduction that comes with such a massive size discrepancy makes this impossible in practice.

As such, a Gaussian Kernel Density Estimator (Gaussian KDE) was used to estimate the PDFs of distributions instead with bandwidth $h$ determined by Scott's Rule (Eq. S3). This provides a non-parametric method of estimating raw observed voxel data PDFs needed for distance calculations while avoiding data loss and artifact introduction from resampling. Here, $h$ represents the determined bandwidth, $n$ represents the number of distribution data points, and $d$is the number of dimensions (1 for univariate data, 2 for bivariate data, etc.).

$$h = n^{-\frac{1}{d+4}} \; (Eq. S3)$$

Once PDFs from Gaussian KDEs have been evaluated, they are then used in Hellinger, Bhattacharyya, and Total Variation distance calculations. Energy and Earth Mover's distance calculations are done via Scipy's built in tools which calculate these measurements directly from distribution data inputs. Hellinger, Bhattacharya, and Total Variation distances were calculated form PDFs by integrating over the PDF bounds as determined below in Equations S4, S5, and S6 using Scipy's quad( ) function from its integration submodule. In these equations, $x$ and $y$ represent the distributions of segmentation ROI and non-segmented liver.

$$Distance = \int_{lb}^{ub} Integrand \;\; (Eq. S4)$$
$$lb = \min([\min(x)\,, \min(y)]) \; (Eq. S5)$$
$$ub = \max([\max(x)\,, \max(y)]) (Eq. S6)$$

**Hellinger Distance**

Hellinger Distance (sometimes known as the Jeffrey's Distance) is used to quantify the similarity between two probability distributions. This is a bounded metric where a maximum value of 1 is achieved when one distribution assigns a probability of 0 to every set in which the second distribution assigns a positive probability, and vice versa (two completely dissimilar distributions achieve the max distance). This distance is symmetrical and is a true metric as non-negativity, the identity of indiscernible, mention symmetry, and the triangle inequality are all satisfied. The equation to solve for Hellinger's distance in the Scipy integral is demonstrated below where $p$ and $q$ represent KDE PDFs for segmented ROI and non-segmented liver.

$$H(p,q) = \frac{\sqrt{\int_{lb}^{ub} (\sqrt{p(x)} - \sqrt{q(x)})^2 dx}}{\sqrt{2}} \; (Eq. S7)$$

**Bhattacharyya Distance**

The Bhattacharyya Distance is a quantity which, like the Hellinger Distance, attempts to quantify the similarity between two probability distributions. For the Bhattacharyya Distance, a value of 0 is achieved when two PDFs being compared are identical. It is based on and related to the Bhattacharyya Coefficient (Eq S7) which represents the overlap between two statistical samples; in Eq S7, $\chi$ represents the discrete domain which PDFs $p$ and $q$ exist on. Like the Hellinger Distance, this is a symmetrical value, but unlike Hellinger Distance, it is not bounded, and it is not a true metric as the triangle inequality is not obeyed. The equation for Bhattacharyya distance in the Scipy integral is given below where $p$ and $q$ represent KDE PDFs for segmented ROI and non-segmented liver.

$$BC(p,q) = \sum_{x \in \chi} \sqrt{p(x)q(x)} \;\; (Eq.S8)$$

$$B(p,q) = -\log\left(\int_{lb}^{ub} \sqrt{p(x)q(x)}dx \;\; (Eq.S9)\right.$$

**Total Variation Distance**

The Total Variation Distance is another distance metric which aims to quantify the difference between two PDFs. Unlike the Hellinger and Bhattacharyya distances which tackle measuring differences in PDFs using more geometric approaches, Total Variation distance (simply) mis defined as half the L1 norm of the difference between two PDFs (the maximum difference between the probability of an event under two distributions). The consequence of this is that it is more sensitive to changes in rare events, meaning that small probabilities can impact this metric more significantly than the two previously discussed distances. This metric is bounded, meaning a value of 0 is achieved when two PDFs are completely dissimilar and 1 is achieved with identical PDFs. Like the Hellinger Distance, it is a true metric which obeys triangle inequality. The equation for Total Variation Distance in the Scipy integral is given below where $p$ and $q$ represent KDE PDFs for segmented ROI and non-segmented liver.

$$TV(p,q) = \frac{1}{2}\int_{lb}^{ub} |p(x) - q(x)|dx \;\; (Eq.S10)$$

**Energy and Earth Mover's Distances**

The Energy and Earth Mover's (sometimes referred to as the Wasserstein's distance) distances did not need to be calculated via manual integration as preexisting functions for these values exist in Scipy (`scipy.stats.energy_distance()` and `scipy.stats.wasserstein_distance()`).

**SHAPE METRIC CALCULATIONS**

Shape metrics were calculated using Pyradiomic's 2D and 3D shape feature capabilities. While not directly related to EPM metrics or other raw MRI voxel intensities, shape features were introduced to the study to determine if automated segmentation ROIs for controls were fundamentally different in geometry from ROIs on true HCC cases. In other words, these metrics aim to test the hypothesis that control ROIs tend to be irregularly shaped noise artifacts that are discernable from more compact, spherical tumor ROIs on cases.

Given that many control (and for that matter, HCC) automated segmentations contain multiple ROIs, a weighting average was implemented which considers the volumes of ROIs when attempting to assign a single shape feature value for multiple ROIs. In determining shape feature values, the largest ROIs (up to 10 maximum) are selected, and the final shape feature value is calculated as shown below; here, $W_n$ is the assigned weight of the $n^{th}$ ROI, $V_n$ is the shape feature value of the $n^{th}$ ROI, where the $n^{th}$ ROI's weight $W_n$ is determined by the ratio of the $n^{th}$ ROI's with the largest ROI volume on a given segmentation mask. This ensures that smaller, less significant features on an automated segmentation do not significantly change shape feature calculations of larger more interesting ROIs, while still being considered proportionally.

$$Shape\ Value = \frac{\sum_{n=1}^{10} W_n V_n}{\sum_{n=1}^{10} Wn} \;\; (Eq.S11)$$

$$W_n = \frac{Vol_n}{Vol_{max}} \;\; (Eq.S12)$$
